# Phrenic Neuropathy Etiologies and Recovery Trajectories in Outpatient Rehabilitation and Neuromuscular Medicine Clinics: A Retrospective Analysis

**DOI:** 10.1101/2024.07.24.24310951

**Authors:** Nicholas Demetriou, Alexandra S. Jensen, Ellen Farr, John M. Coleman, Senda Ajroud-Driss, Adenike A. Adewuyi, Lisa F. Wolfe, Colin K. Franz

**Affiliations:** Feinberg School of Medicine, Northwestern University, Chicago, IL; Shirley Ryan AbilityLab (formerly the Rehabilitation Institute of Chicago), Chicago IL; McGaw Medical Center of Northwestern University, Chicago, IL; Pulmonary and Critical Care, Department of Medicine, Northwestern University, Chicago, IL; Ken and Ruth Davee Department of Neurology, Northwestern University, Chicago, IL; Physical Medicine and Rehabilitation, Northwestern University, Chicago, IL; Kimberly K. Querrey and Louis A. Simpson Institute for Bioelectronics, Northwestern University, Chicago, IL

## Abstract

**Introduction/Aims:** Phrenic Neuropathy (PhN) impairs diaphragm muscle function, causing a spectrum of breathing disability. PhN etiologies and their natural history are ill defined. This knowledge gap hinders informed prognosis and management decisions. This study aims to help fill this knowledge gap on PhN etiologies, outcomes, and recovery patterns, especially in the context of non-surgical clinical practice.

**Methods:** This is a retrospective study from two interdisciplinary clinics, physiatry and neurology based. Patients were included if PhN was identified, and other causes of hemi-diaphragm muscle dysfunction excluded. Patients were followed serially per the discretion of the neuromuscular trained neurologist or physiatrist. Recovery was assessed using pulmonary function tests (PFTs), diaphragm muscle US thickening ratio, and patient-reported outcomes in patients presenting within two years of PhN onset.

**Results:** We identified 151 patients with PhN. The most common etiologies included idiopathic (27%), associated with cardiothoracic procedure (24%), and intensive care unit (17%). Of these patients, 117 (77%) were evaluated within two years of PhN onset. Of patients included in outcome analyses, 69% saw improvement on serial US, 50% on serial PFTs and 79% reported symptomatic improvement at an average of 13, 16, and 17 months respectively.

**Conclusion:** This study maps PhN etiologies and recovery. A clear majority of PhN patients show improvement in diaphragm muscle function, but on average improvements took 13-17 months depending on the assessment type. These insights are vital for developing tailored treatments and can guide physicians in prognosis and decision-making, especially if more invasive interventions are being considered.

## Introduction

Phrenic neuropathy (PhN) is a debilitating condition characterized by the impairment of the diaphragm muscle, the primary inspiratory muscle. This condition poses significant challenges to affected individuals, impairing their respiratory function and reducing quality of life.

Despite its profound consequences, the true incidence and prevalence of PhN are ill-defined with a specific lack of literature collating data across the various PhN etiologies. Prior studies tend to shed light on specific subsets of PhN etiologies and imply we may have previously underestimated the scale of the problem. PhN has been observed as a complication in various medical contexts, affecting an estimated 1-2% of cardiac surgery patients^1, 2^, 3.2%-29.6% of lung transplant recipients^3-6^, 3.2% of patients hospitalized with COVID-19^7^ and 6-14% of patients with neuralgic amyotrophy (NA)^8-10^. However, there is a much longer list of potential causes of PhN that have been reported^11^. Importantly, PhN and its sequelae are compounded by other common health diagnoses such as hypertension^12^, diabetes mellitus^13, 14^, various lung diseases^15^, cancer^16^, and immunosuppression^17^. Altogether this highlights the variable etiological origins of PhN and the potential scale of PhN as a public health concern.

Diaphragm muscle ultrasound (US) has provided clinicians with a non-invasive tool that appears to offer both high sensitivity and specificity for diagnosis^18^, challenging electrodiagnostic studies as the gold standard for assessing phrenic nerve health^19^. PhN is commonly diagnosed via diaphragm muscle US or fluoroscopy (sniff test) with paradoxical or significantly decreased diaphragm muscle movement^20^. However, despite the wide range of available diagnostics, significant knowledge gaps persist in the literature regarding the natural history and expected recovery trajectories of PhN patients, particularly those managed non-operatively.

PhN management and prognosis are dependent on the etiology and significance of respiratory impairment. Patients with mild respiratory dysfunction may be managed conservatively with optimization of concurrent medical conditions and inspiratory muscle training^21^. Those with significant respiratory impairment may be offered interventions ranging from nocturnal non-invasive ventilation to surgical interventions targeting the phrenic nerve or diaphragm muscle. Due to limited data on natural history of PhN and lack of standardization on tracking PhN recovery, treating physicians can at best draw upon more generic principles^22^ of peripheral nerve injuries in making prognostic and treatment decisions (e.g. axons regenerate about 1 mm/day). Importantly, most PhN specific outcome studies to date focus on post-intervention cases, primarily involving surgical procedures such as phrenic reconstruction^23-25^ and diaphragm muscle plication^26-28^. As a result, the current literature lacks high-quality data to support surgical versus non-surgical management, thus making it difficult to advise on such interventions.

This study has a dual objective: firstly, to elucidate the distribution of PhN etiologies, shedding light on the diverse origins of this condition with specific focus on how it presents itself to interdisciplinary outpatient clinics based in neurology and physiatry neuromuscular practice; and secondly, to define the recovery potential and trajectories of patients with PhN.

## Methods

### Subjects

This is a retrospective study which assessed patients at two interdisciplinary clinics including neuromuscular medicine (neurologist or physiatrist) and pulmonology physicians at either Northwestern Memorial Hospital or the Shirley Ryan AbilityLab both located in Chicago, IL (USA). Patients were referred to this clinic with symptomatic PhN often initially suspected by hemi-diaphragm muscle elevation on radiographic imaging. Patients were frequently evaluated via point of care diaphragm muscle US. The final diagnosis of PhN was by at least two physicians (a board-certified pulmonologist and a board-certified physiatrist or neurologist subspecialized in neuromuscular medicine) on the basis of clinical history, physical exam, and the aggregation of available test results including phrenic NCS, chest radiographs, fluoroscopy, and pulmonary function tests (PFTs). Diaphragm muscle US, used to assess the vast majority of patients during their initial presentation, was obtained through diagnostic study performed by a musculoskeletal radiology or a neuromuscular medicine certified physiatrist with certificate in neuromuscular ultrasound (C.K.F.). Diaphragm muscle US was performed based on the protocol described by Boon and colleagues^29^. Briefly, patients were seated in a reclined position (∼45 degrees) with the probe placed at the 8th or 9th intercostal space in the midaxillary line. Diaphragm muscle thickness was then measured at the end of full inspiration and normal expiration. Serial diaphragm muscle US was recommended at 3-to-6-month intervals dependent on the patient’s clinical status and ability to follow up (e.g. local versus out of state patients). Many patients were also assessed with serial PFTs performed either in outpatient PFT laboratory or in office subsequently interpreted by the attending pulmonologist (L.F.W. or J.M.C.). PFTs were reported as the best of three trials spaced each by one minute. The interval for serial PFT testing was left to the discretion of a treating physician on the interdisciplinary team but was noted to be more challenging during this period due to COVID-19 related restrictions on aerosol generating office procedures.

Diaphragm muscle dysfunction secondary to other conditions which may affect phrenic or diaphragm function, including stroke, spinal cord injuries, ALS, Guillain-Barre syndrome, myasthenic syndromes, myositis, and muscular dystrophies, were excluded from this cohort as such illnesses blur the cause of the patient’s diaphragmatic dysfunction^30^. Furthermore, patients initially suspected of having PhN and later determined to have a progressive degenerative disease, such as ALS, were excluded from this study. One patient, diagnosed with PhN decades prior to their IBM diagnosis, was included in our study, however, they were excluded from outcome analyses since they were a chronic PhN patient.

Patient subjective reports of respiratory symptoms in the electronic medical record were also employed to assess the onset and recovery of PhN. A subset of patients (N=117) was assessed longitudinally to determine improvement in phrenic, diaphragm muscle, and respiratory function. Patients with chronic PhN, defined as having their initial PhN evaluation two or more years after their injury onset, were excluded from outcome analyses as spontaneous recovery beyond 2 years for peripheral nerve injury is very unlikely^31^. Prior surgical intervention, offered to a small fraction of our cohort, was tracked, however, this was not an exclusion criterion.

### Measures

Patient data was collected longitudinally through serial clinic visits and patient medical charts. Patient outcomes were assessed using serial diaphragm muscle US scans, PFTs and documented patient subjective outcomes. To assess diaphragm muscle function via US, we used the thickening ratio (TR), calculated as diaphragm muscle thickness at the end of a full inspiration (total lung capacity) divided by the diaphragm muscle thickness at the end of a normal expiration (functional residual capacity). A thickening ratio of less than 1.2 indicates diaphragm muscle dysfunction^32^. In our patient outcomes analysis, individuals were categorized as improved if a subsequent diaphragm muscle US demonstrated a TR of 1.2 or greater.

Patient respiratory function and recovery was further assessed via serial PFTs. Prior studies have concluded that an absolute change of 5% in FVC from baseline was a substantial and significant change in respiratory function^33^. This cut-off was applied to our cohort to designate individuals as improving (>5% increase in FVC) or not improving in their respiratory function (<5% increase in FVC). Our third outcome measure assessed patient improvement qualitatively through patient interviews. To collect this qualitative data, all notes from the interdisciplinary clinic were reviewed for mentions of patient reported respiratory improvement vs no improvement. Finally, we collated all three outcome measures at the patient level to create a combined view of outcomes to identify patients who exhibited improvement in at least one of these outcome measures.

### Analysis

Sample characteristics were calculated using percentages and means with standard deviations. Analysis employs descriptive statistics to highlight frequency of different PhN etiologies and their recovery rates.

## Results

In total 151 patients (table 1) screened at our outpatient clinic were identified as having a PhN assessed via diagnostic tests listed in table 2. Patient ages at the time of PhN onset ranged from 2 to 88 years old with a mean of 58 and median of 63. In total nine patients in our sample received a surgical phrenic nerve intervention, either diaphragm muscle plication (n=6) or diaphragm muscle pacer (n=3). Five of these patients had chronic PhN injuries at their initial presentation and the remaining four were excluded from outcomes analysis. In addition, 19 non-chronic PhN patients received medication related treatment in the form of immune and inflammation suppression, predominantly prednisone, however two patients also received IVIG, and one received plasma exchange. Medical treatment was primarily geared towards those with NA and idiopathic PhN if clinical concern for dysimmune pathology. Thirteen patients with non-chronic PhN passed away within two years of their PhN onset, seemingly for reasons not directly related to their PhN, three of whom have reported outcomes.

**Table 1:** Sample Characteristics.

| Subject Characteristics | Sample<br>Values<br>(n=151) | Cardio-<br>thoracic<br>(n=36) | ICU (n=26) | Idiopathic<br>(n=41) | NA (n=31) | Trauma<br>(n=6) | Tumor/Mas<br>s (n=11) |
| --- | --- | --- | --- | --- | --- | --- | --- |
| Age (years), Mean (SD) | 58 (15) | 56 (19) | 54 (14) | 67 (11) | 57 (12) | 40 (19) | 55 (15) |
| Female Sex, N (%) | 59 (39%) | 12 (33%) | 10 (38%) | 10 (49%) | 11 (35%) | 0 (0%) | 6 (55%) |
| BMI (kg/m²), Mean (SD) | 30 (7) | 27 (6) | 31 (10) | 31 (6) | 32 (7) | 23 (7) | 28 (6) |
| Comorbidities, N (%) |  |  |  |  |  |  |  |
| Hypertension | 91 (61%) | 23 (64%) | 19 (73%) | 31 (76%) | 13 (42%) | 1 (17%) | 4 (36%) |
| Diabetes/ Pre-Diabetes | 79 (53%) | 20 (56%) | 18 (69%) | 24 (59%) | 13 (42%) | 1 (17%) | 3 (27%) |
| Lung Disease<br>(COPD, ILD, etc) | 55 (36%) | 18 (50%) | 12 (46%) | 15 (37%) | 7 (23%) | 1 (17%) | 2 (18%) |
| Cancer | 29 (19%) | 7 (19%) | 2 (8%) | 7 (17%) | 4 (13%) | 1 (17%) | 8 (73%) |
| Transplant/ Immune<br>Compromised | 27 (18%) | 17 (47%) | 6 (23%) | 1 (2%) | 0 (0%) | 1 (17%) | 2 (18%) |

**Table 2:**
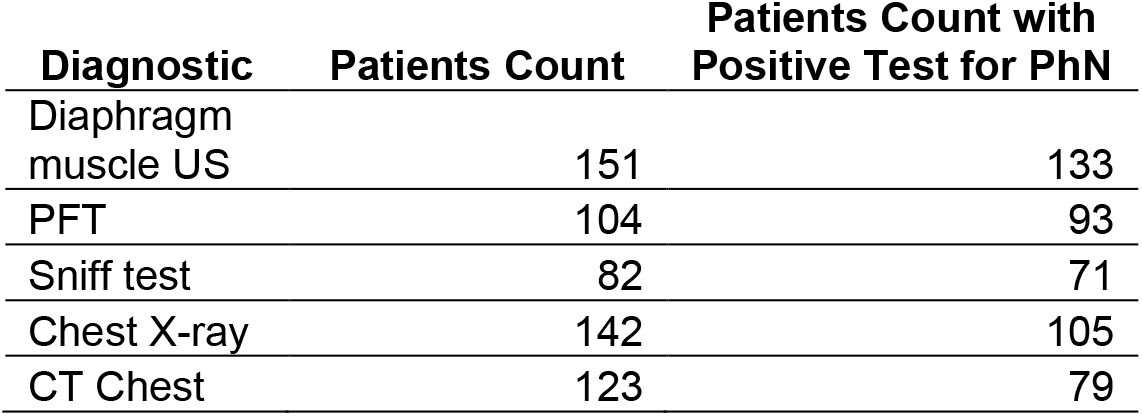
Diagnostic Tests of Cohort.

| Diagnostic | Patients Count | Patients Count with<br>Positive Test for PhN |
| --- | --- | --- |
| Diaphragm<br>muscle US | 151 | 133 |
| PFT | 104 | 93 |
| Sniff test | 82 | 71 |
| Chest X-ray | 142 | 105 |
| CT Chest | 123 | 79 |

Of the 117 non-chronic (i.e. initial assessment date was <2 years from onset) PhN patients, all received diaphragm muscle US screening, with 106 (91%) demonstrating abnormal TR on initial presentation. Additionally, 78 patients had at least one set of PFTs completed, 69 (88%) with an abnormally low FVC (FVC < 80% predicted)^34^. Diaphragm muscle US outcomes were assessed in 62 patients, serial PFTs in 46 patients (30 with both diaphragm muscle US and PFTs) and a self-reported outcome in 48 patients. When collating all three outcomes at the patient level, 86 patients had at least one reported outcome measure. Considering all individuals with serial diaphragm muscle US, 69% saw improvement in their diaphragm muscle function (TR>1.2). Of the 46 individuals with serial PFTs, 50% saw improvement (FVC Change >5%). Of the 48 patients with a self-reported outcome, 79% reported their status as improved. Of the 86 patients with at least one reported outcome measure, 66% exhibited improvement on at least one measure. A breakdown of improvement by outcome measure and phrenic nerve injury etiology can be found in figures 1A-1D. For patients with serial diaphragm muscle US scans (≥3), an increase in diaphragm muscle thickness preceded improvement in diaphragm muscle thickening ratio (TR) 38% of the time, with nearly all remaining patients in this subgroup seeing both increased diaphragm muscle thickness and TR at the time of initial improvement.

**Figure 1:**
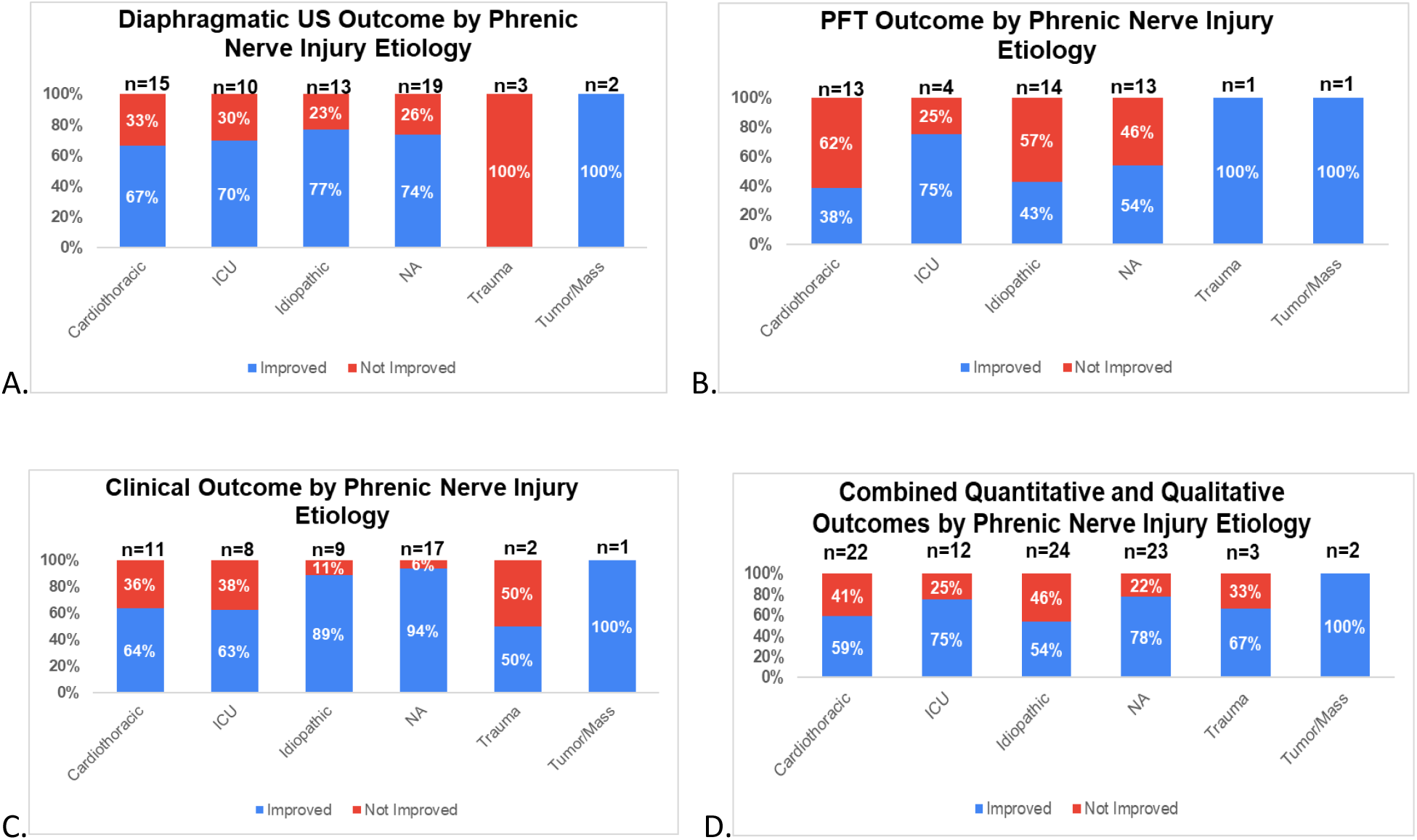
Percentage of patients achieving improvement in serial outcome measures: (A) US, (B) PFTs, (C) Subjective clinical (D) Combined outcome measures

Phrenic NCS were completed in 79 non-chronic PhN patients as part of their work up. Of these, 6 patients were identified to have normal response, 9 had an impaired response, and the remaining (n=64) had no response (concerning for complete axonotmesis). Of the 9 patients with incomplete axonotmesis, 4 had serial diaphragm muscle US, 2 (50%) demonstrating improvement. One (100%) had serial PFTs with noted improvement, and 5 of 5 (100%) with a reported clinical outcome, reported subjective improvement.

Mean time to improvement (figure 2) varied by outcome measure. On average, it took 13 months to identify ultrasonographic diaphragm muscle improvement with a range from 1 to 44 months. Using serial PFT’s, it took on average 16 months to identify improvement in respiratory capacity, with a range from 2 to 35 months. Clinically (subjective) reported recoveries occurred on average 17 months following PhN with a range of 2-42 months.

**Figure 2:**
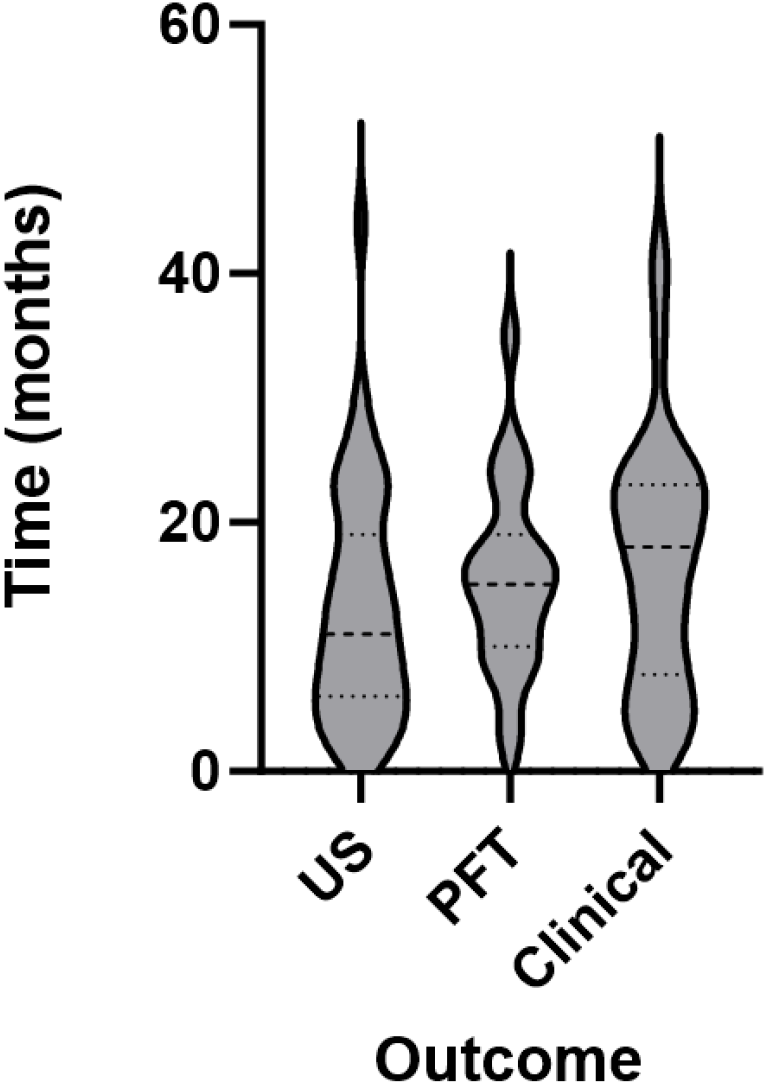
Distribution of recovery times as measured via diaphragm muscle US, PFTs, and self-reported improvement.

## Discussion

The etiological distribution of PhN at our institutions was led by idiopathic, cardiothoracic procedure related, NA and ICU. This contrasts with earlier studies which described cardiothoracic/trauma as the primary cause of PhN paralysis^35^. To date, most prior studies have focused on a single specific PhN etiology (e.g. post-lung transplant). Thus, our study stands out due to the relative diversity of PhN etiologies encountered as well as our emphasis on conservative management (i.e. <6% of our patients had a surgical intervention – plication or diaphragm pacer implant).

Abnormalities on diaphragm muscle US and PFTs proved very common among patients with PhN. A small minority of patients had normal range values for these tests at initial presentation, however, their presentation and prior tests (e.g. sniff test, electrodiagnostic studies) were consistent with PhN. A smaller percentage of patients received PFTs compared with diaphragmatic ultrasound likely due limitations during the Covid-19 pandemic and because PFTs often required an additional healthcare visit while diaphragmatic ultrasound was completed during clinic visits. Comparing recovery based on diaphragmatic US response to PFT improvement found moderately increased recovery rates on diaphragm muscle US. In addition, our results suggest that increased diaphragm muscle thickness may be predictive of early recovery. Thus, regular diaphragm muscle US may aid physicians in developing prognosis trajectories for patients. Interestingly, a notably larger percentage of patients experienced symptomatic improvement compared with those who improved on US or PFTs. Importantly, many factors, such as improved training of accessory breathing muscles, weight loss, and comorbidity status may impact a patient’s breathing ability over time and compensate for a person’s PhN. This suggests the potential value of interdisciplinary, non-surgical focused care for patients with PhN to provide patient education, and to help direct care including nocturnal non-invasive ventilation and rehabilitation services.

Our data shows that patients with PhN secondary to NA have one of the highest rates of improvement among PhN etiologies and tracks closely to recovery rates seen in NA in general^36^. NA was diagnosed in patients with concurrent brachial plexopathy or clinical features highly suggestive of NA such as intensely painful onset, usually in shoulder/periscapular region, with patchy areas of paresis, pain or sensory deficits^37^. The reason for the relatively high recovery rate across NA patients is unclear, however it may relate to the pathophysiology of the underlying nerve injury. NA can lead to either a demyelinating (more rapid recovery possible) or axonal nerve injury (slower recovery potential) while iatrogenic causes are predominantly axonal^38^.

Interestingly those with idiopathic and cardiothoracic procedure related injuries showed lower recovery rates across collated outcomes at approximately 50% for each. Patients with a cardiothoracic procedure related and idiopathic PhN etiology had some of the highest comorbidity burden, which may have impacted their recovery patterns. Also, lack of understanding of the mechanism of injury in the idiopathic PhN group challenges our ability to explain this result and is an opportunity for future research. Further research may reclassify these individuals and provide better insight as to why they have relatively poorer outcomes. Our data on PhN recovery rates from cardiothoracic procedure associated origin is incongruent with a prior study which reported a near complete recovery rate^39^. However, compared to this earlier study, the degree of nerve injury in our study populations was likely significantly greater. This is expected, due to numerous factors. First off, the referral nature of our clinic site indicates patients’ PhN were symptomatic and relatively long standing, in comparison to this prior prospective study which included all cardiothoracic surgical patients regardless of symptom status. Furthermore, most patients in this earlier trial had intact NCS following surgery, whereas our study population, for the most part, did not.

Late, spontaneous recovery in a majority of people with PhN which was demonstrated within this case series and other case series that emphasized conservative management^40^ highlights that conservative management can be quite effective and may be the preferred first line PhN treatment for many patients. No prospective, randomized head-to-head comparisons exist between conservative and invasive treatments options such as diaphragm plication surgery. Several conservative interventions may improve a patient’s symptoms, but more research is needed to establish evidence-based treatment guidelines. Two such conservative interventions include respiratory muscle training and non-invasive ventilation^41-43^.

The present outcomes after conservative management of PhN calls into question the optimal timing and role for phrenic nerve reconstruction. It is difficult to compare conservative management of PhN against interventional case series that offer various phrenic nerve repair to PhN patients^44^. If the PhN is known to involve neurotmesis (nerve is completely severed) consideration of phrenic repair (grafting, nerve transfer) is reasonable given positive results seen in select populations^45^. Interestingly, the vast majority of the patients in our study likely exhibited an axonotmesis pattern (i.e. nerve structure, but not axons remain in continuity), since they did not have penetrating or high force nerve injury mechanism, but did have an absence of an evoked phrenic motor response, yet the majority of them saw improvement without surgery. Our sample size of patients with incomplete/partial axonotmesis is too small to draw strong conclusions from, however, they seem to show similar improvement rates when compared with the whole group with particularly strong subjective improvement. Given that nerve reconstruction surgery is a time sensitive intervention, the role it should play for most PhN patients remains uncertain since so many will improve over time without surgery. In addition, if patients do not recover diaphragm muscle function to an adequate extent following conservative treatment for >2 years, diaphragm muscle plication remains an option that can offer comparable rates of benefit^46^.

Limitations of this study stem from its nature as a retrospective cohort study. Patient outcome data was tracked at variable intervals and likely overestimated timeframes needed for patient recoveries in this study. Furthermore, patient reported outcomes are prone to recall bias. Additionally, the reliance on existing data, which may be imperfect or incomplete and is dependent on patient follow up, opens the possibility for reporting bias. Thus, extrapolation of the results presented here is limited and should be conducted cautiously.

## Conclusion

In conclusion, this study sheds light on the diverse etiologies of PhN, emphasizing the complexity of this condition beyond traditional cardiothoracic and trauma-related causes. Our findings reveal that PhN, while challenging to diagnose and manage, offers some promise of spontaneous recovery. The variability of recovery rates between different PhN etiologies suggests that the underlying mechanisms can inform the recovery process. This research marks a step toward understanding the complexities of PhN and lays a path for future prospective trials, aiming to refine treatment strategies and enhance patient outcomes in the realm of PhN.

## Data Availability

All data produced in the present study are available upon reasonable request to the authors.

## Acknowledgements

C.K.F. would like to acknowledge support from the Belle Carnell Regenerative Neurorehabilitation fund.

## Abbreviations

FVC: Forced Vital Capacity
NA: Neuralgic amyotrophy
PFT: Pulmonary function test
PhN: Phrenic Neuropathy
TR: Thickening Ratio
US: Ultrasound

